# Impact of voluntary risk-mitigation behaviour on transmission of the Omicron SARS-CoV-2 variant in England

**DOI:** 10.1101/2022.01.26.22269540

**Authors:** Ellen Brooks-Pollock, Kate Northstone, Lorenzo Pellis, Francesca Scarabel, Amy Thomas, Emily Nixon, David A. Matthews, Vicky Bowyer, Maria Paz Garcia, Claire J. Steves, Nicholas J. Timpson, Leon Danon

## Abstract

**Background:** The Omicron variant of SARS-CoV-2 infection poses substantial challenges to public health. In England, “plan B” mitigation measures were introduced in December 2021 including increased home working and face coverings in shops, but stopped short of restrictions on social contacts. The impact of voluntary risk mitigation behaviours on future SARS-CoV-2 burden is unknown.

**Methods:** We developed a rapid online survey of risk mitigation behaviours during the winter 2021 festive period and deployed in two longitudinal cohort studies in the UK (Avon Longitudinal Study of Parents and Children (ALSPAC) and TwinsUK/Covid Symptom Study (CSS) Biobank) in December 2021. Using an individual-based, probabilistic model of COVID-19 transmission between social contacts with SARS-CoV-2 Omicron variant parameters and realistic vaccine coverage in England, we describe the potential impact of the SARS-CoV-2 Omicron wave in England in terms of the effective reproduction number and cumulative infections, hospital admissions and deaths. Using survey results, we estimated in real-time the impact of voluntary risk mitigation behaviours on the Omicron wave in England, if implemented for the entire epidemic wave.

**Results:** Over 95% of survey respondents (N_ALSPAC_=2,686 and N_Twins_=6,155) reported some risk mitigation behaviours, with vaccination and using home testing kits reported most frequently. Less than half of those respondents reported that their behaviour was due to “plan B”. We estimate that without risk mitigation behaviours, the Omicron variant is consistent with an effective reproduction number between 2.5 and 3.5. Due to the reduced vaccine effectiveness against infection with the Omicron variant, our modelled estimates suggest that between 55% and 60% of the English population could be infected during the current wave, translating into between 15,000 and 46,000 cumulative deaths, depending on assumptions about vaccine effectiveness. We estimate that voluntary risk reduction measures could reduce the effective reproduction number to between 1.8 and 2.2 and reduce the cumulative number of deaths by up to 24%.

**Conclusions:** We conclude that voluntary measures substantially reduce the projected impact of the SARS-CoV-2 Omicron variant, but that voluntary measures alone would be unlikely to completely control transmission.

## Introduction

Throughout the COVID-19 pandemic, human behaviour has modified and controlled transmission. In the initial epidemic growth phase in early 2020, government-defined social distancing, including school closures and stay-at-home orders, was widely used worldwide to prevent the exponential growth in numbers of cases. However, as the pandemic progressed and personal biosecurity measures have become available, the role of individual choice in using face coverings, rapid home tests and vaccination has become increasingly important.

A challenge for predicting future SARS-CoV-2 burden is accurately capturing the population response to measures and advice. The first impositions of stay-at-home orders were widely adhered to[1,2]. Behaviours changed rapidly upon the imposition of restrictions, but were slower to revert to ‘normal’ once restrictions were lifted, with a high proportion of individuals still practicing biosecurity measures such as wearing face coverings and limiting travel[3]. Such behaviours are influenced by multiple factors including trust in the government, individual perception of risk, media reporting and personal interactions, and the importance of different factors varies over time[4].

Behavioural dynamics, which affect both transmission potential and reporting of cases[5], are increasing included in infectious disease models, although with varying degrees of detail. The availability of Google Mobility data [3] during the COVID-19 pandemic has been used as a proxy for changing mobility and contact behaviour in transmission models[6–8], but it is necessarily retrospective in nature and assumptions about future behaviour are required. Direct, real-time measures of behaviour, such as social contact surveys[1,9], are limited.

Here, we report on the results of a rapid, prospective survey of intended behaviours over the 2021 winter festive period. The purpose of the study was to estimate the impact of voluntary behaviours on SARS-CoV-2 transmission and numbers of cases and deaths due to the Omicron SARS-CoV-2 variant, first identified in November 2021. In the UK at the time there were minimal legal restrictions on behaviours, although due to the Omicron variant, the UK government announced a ‘plan B’ on 8 December 2021, in which face masks were compulsory in indoor public spaces and home working was encouraged.

At the time this work was undertaken in December 2021, there was still considerable uncertainty about the transmission potential and severity of the Omicron variant compared to the previously dominant Delta variant. The rapid growth, with cases due to Omicron doubling every 2 to 3 days, suggested a substantial advantage over other variants [10,11]. This transmission advantage could be due to an increase in transmission potential and/or immune escape mutations which render immunity from vaccination and previous infections less protective against infection than before [12,13]. Emerging evidence suggests that infection with Omicron is less likely to lead to severe disease and death[14,15], although with considerable uncertainty.

## Methods

### Ethics statement

Ethical approval for the study was given by the Avon Longitudinal Study of Parents and Children (ALSPAC) Ethics and Law Committee on 25 November 2021. Participation was voluntary and only anonymised data were collected. The survey was sent out to TwinsUK participants under existing TwinsUK ethics (REC reference: EC04/015) and TwinsUK BioBank ethics (REC reference: 19/NW/0187), granted by the North West - Liverpool Central NHS Research Ethics Committee (England) (https://www.hra.nhs.uk/planning-and-improving-research/application-summaries/research-summaries/twinsuk-biobank/). The survey was sent out to COVID Symptom Study (CSS) Biobank participants under existing CSS Biobank ethics (REC reference: 20/YH/0298), which was granted by the Yorkshire & The Humber - Leeds East NHS Research Ethics Committee (England) (https://www.hra.nhs.uk/planning-and-improving-research/application-summaries/research-summaries/covid-symptom-study-biobank-covid-19/).

### Rapid survey of social contacts and risk-mitigation behaviour during December 2021

We developed an online survey about plans for Christmas 2021 to fill the gap in social contact and behavioural data. The survey covered the festive period from 20 December 2021 to 2 January 2022 and included questions about planned face-to-face interactions, numbers of households meeting indoors, vaccination and risk-mitigation behaviours (complete list of questions in the Supplementary Information). The survey was advertised to the participants of three longitudinal cohorts: the Avon Longitudinal Study of Parents and Children (ALSPAC)[16– 18], TwinsUK [19,20] and the COVID Symptom Survey (CSS) Biobank [21]. TwinsUK and the CSS Biobank were managed by the same team and were treated as one combined cohort for this study.

ALSPAC is an intergenerational prospective birth cohort from the southwest of England. The study recruited 14,541 pregnant women with expected dates of delivery between 1^st^ April 1991 to 31^st^ December 1992 in the former county of Avon and has followed the women, their partners and children since. Full details of the cohort and study design have been described previously[16–18] and are available at www.bristol.ac.uk/alspac. The ALSPAC survey was deployed using Microsoft Forms. The survey was an anonymous, standalone survey, and data were not linked to any other data on participants. The survey link went live on 9 December 2021 and was active until 22 December 2021. Participants of ALSPAC were invited to participate via a link in the annual newsletter which went to participants on 15 December 2021 and via social media posts, although anyone with the link could complete the survey.

TwinsUK is a UK registry of volunteer twins in the United Kingdom, with about 14,000 registered twins[19,20]. The Covid Symptom Study (CSS) Biobank is a longitudinal study run by researchers at King’s College London with approximately 12,000 participants [21]. The TwinsUK/CSS Biobank survey was implemented in REDCap, accessible via an anonymous link advertised in the Christmas newsletter. The survey link was active from 15 to 20 December 2021.

Data from the surveys were analysed in R version 4.01. We calculated descriptive statistics by age and used a binomial general linear model to explore associations between risk mitigation behaviours.

### Modelling approach

We used an individual-based disease model based on social contact data, validated against the early growth of SARS-CoV-2 in England in 2020 [22]. The basic premise behind the approach is that we calculate a distribution of individual reproduction numbers for the entire population, based on individuals’ reported social contacts. Say individual *i* has *k*_*i*_social contacts on a given day. Each *k*th social contact involves *n*_*k*_ other individuals and it lasts for a time *d*_*k*_, which acts to weight the number of contacts. Their personal individual reproduction number (i.e. the number of secondary cases they generate) is given by

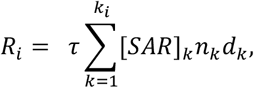

where [*SAR*]_*k*_ is the Omicron-specific secondary attack rate (proportion of contacts that result in secondary infection) for the setting of the social contact, either household or non-household and *τ* is a constant calibrated to the reproduction number *R*_0_ = 7 for the Delta variant (i.e. with Delta-specific secondary attack rates) in the absence of vaccination or natural immunity. We used social contact data from the Social Contact Survey (SCS)[23] and secondary attack rates estimated by UKHSA from positive tests in contacts named to NHS Test and Trace [14]. We use [*SAR*]_*k*_ to weight the type of contact (e.g., household contacts are more “intimate” than non-household ones) but assume that, for a given type of contact, all individuals have the same infectivity and susceptibility.

To calculate the population-level reproduction number from the individual reproduction numbers, we assume proportionate mixing between individuals, i.e. that the probability of contacting individual *j* is proportional to their number of contacts over the total number of contacts in the population, *R*_*j*_/∑_*j*_ *R*_*j*_. The population-level reproduction number therefore scales with the square of the individual-level reproduction numbers:

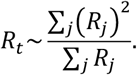

We use the individual reproduction numbers to calculate the cumulative numbers of cases, hospital admissions and deaths. We use the notation *σ*_*j*_ to denote the probability that individual *j* does not get infected during the ensuing epidemic wave. *σ*_*j*_ depends on the susceptibility of individual *j* but also on the infectiousness of all other individuals, and the probability that they do/do not get infected, therefore there is no closed form solution for calculating *σ*_*j*_. Following [24], *σ*_*j*_ can be shown to be

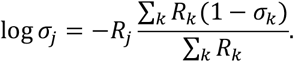

As there is no closed form solution for calculating *σ*_*j*_, it is calculated by iteration, starting with *σ*_*j*_ = 0.5 for all *j*s and recalculating all final sizes, repeating until the estimates converge.

The cumulative number of cases is calculated from the individual *σ*_*j*_s by multiplying by an individual-specific weight *w*_*j*_ based on the representativeness of the social contact survey that is used for the model. The total number of cases is ∑_*j*_ *w*_*j*_(1 − *σ*_*j*_). The number of deaths is calculated from the number of cases, multiplied by the age-specific infection fatality rate, ∑_*j*_ *w*_*j*_(1 − *σ*_*j*_)*μ*_*j*_.

### Modelling vaccination and natural immunity

We capture vaccination using the vaccination line list data provided by UKHSA on 26 November 2021. We aggregated the data to calculate the proportion by age that had received a single or double dose of each of the main available vaccines in England (AstraZeneca or Pfizer/Moderna), and a booster dose, leading to five categories. We estimated the proportion of individuals by age with immunity from a natural infection using the Pillar 2 data of test positive cases, assuming that 50% of infections were identified as cases. Therefore, we model the population using eight categories of vaccine/immune status: five vaccination states, immunity from vaccination and a natural infection, immunity from natural infection only and no immunity/unprotected. We made the simplifying assumption that vaccine and infection status were independent – the proportions of each age group falling within the eight categories were calculated using data from 26 November 2021 (supplementary table 1).

The effect of vaccination is incorporated into the model via three mechanisms: by reducing the probability that an individual is infected (reduced susceptibility), reducing the probability that the individual will transmit to others (reduced transmissibility) and reducing the risk of severe disease and death. We use UKHSA estimates of vaccine effectiveness for the Delta variant [15] and scale the effectiveness against infection and transmission down by squaring the effectiveness expressed as a proportion – therefore an effectiveness of 80% against Delta becomes 64% against Omicron [25].

The individual reproduction number is modified by vaccination by reducing the probability of transmission

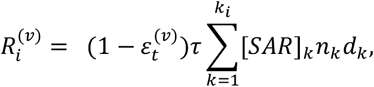

where 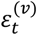 is the vaccine effectiveness against transmission for vaccine/immunity state *v*.

The population-level reproduction number is formed of all vaccine states

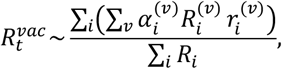

where 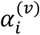 is the probability that individual *i*is in vaccination state *v*, such that 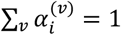 and 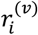 is the ‘receiving’ risk of infection,

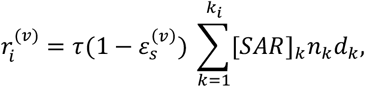

with 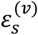 being the vaccine effectiveness against infection for vaccine state *v* and *k* a constant calibrated to the initial reproduction number without vaccination.

The final size calculations are also modified by the action of the vaccine,

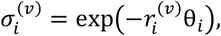

Where

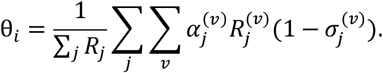

The cumulative number of cases is calculated as

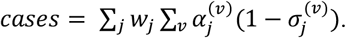

The cumulative number of hospital admissions is calculated with the individual-specific hospital admission rate *h*_*j*_ as:

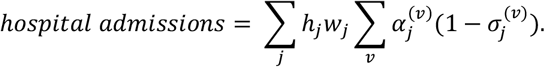

Finally, the cumulative number of deaths is calculated using the individual-specific mortality admission rate *μ*_*j*_ as:

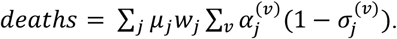

### Risk mitigation measures

This framework allows us to model risk mitigation measures at an individual level. In the model, an individual is associated with a probability of exhibiting risk-mitigation behaviours, according to age, and determined by survey responses. For each model iteration, an individual is determined to practice that risk mitigation measure or not, based on a random number draw. For example, in persons aged 30-39: 67% report limiting in-person visits to shops, 59% report using a face mask, 51% report avoiding public transport, 47% report working from home and 81% report using home testing kits. So, for a single model run for an individual aged 35, we draw a random number, say *rand*_1_ = 0.69, in which case, this individual would not limit visits to shops or public transport use, or use a face mask, or work from home, but would use home testing kits. We assume a single number determines all risk mitigation measures for an individual, rather than choosing each of them independently, reflecting the evidence that some individuals tend to be more cautious than others in all their activities.

Contact tracing is applied to symptomatic cases only (where the probability of symptoms is determined by the age of the individual), implemented by reducing the number of secondary cases by a proportion *CTF*, determined from the NHS Test and Trace statistics as approximately (proportion of cases reached and asked to provide details of recent close contacts)x(proportion who provided details for one or more close contact)x(Proportion of contacts reached within 24 hours)x(proportion of close contacts reached and asked to self-isolate)[26].

Lateral flow testing of asymptomatic cases is implemented in a similar way to contact tracing but originating from asymptomatic cases. Individuals were given an age-specific probability of using home testing kits based on the results of the ALSPAC survey. If a home testing kit was used and infection was identified, the number of secondary cases is reduced by the same proportion as for contact tracing, *CTF*. The sensitivity of lateral flow testing was taken as *s*_*L*_ = 50%[27].

Mask wearing was implemented by reducing the probability of transmission by a proportion *CS*. Our estimate for the current impact of mask-wearing on transmission is less than 25%. In a 2020 systematic review, Chu et al. reported a smaller risk reduction for face mask use in non-healthcare settings compared to for healthcare settings, and a smaller reduction for single layer face masks as opposed to respirators and surgical masks [28,29].

Working from home, limiting in-person shopping, and avoiding public transport were implemented by eliminating contacts reported as occurring at work/in shops/on public transport. If a contact did not take place, we set *n*_*k*_ = 0 for that interaction. In addition, we simulated school holidays over Christmas by removing all contacts for children under 18-years-old with “school” listed as the context, as these were assumed not to take place during the winter holidays.

### Changes to disease severity

We investigate three main severity scenarios for the Omicron variant: a) a 40% reduction in mortality rates associated with Omicron infection and a reduction in vaccine effectiveness against severe disease compared to the Delta variant; b) a 20% reduction in mortality rates associated with Omicron infection and a reduction in vaccine effectiveness against severe disease compared to the Delta variant; c) a 20% reduction in mortality rates associated with Omicron infection with no reduction in vaccine effectiveness against severe disease.

### Model implementation

The model was written in R version 4.01. The population of individuals was simulated 10 times and results aggregated. A summary of parameter values and interpretations is given in Table 1. The model code is available at https://github.com/ldanon/ALSPAC_CO_Survey.

**Table 1:** Parameter values used in the model.

| Parameter name/interpretation/symbol |  | Parameter estimates/ranges and sources |
| --- | --- | --- |
| Secondary Attack Rate, [SAR] | Delta, household | 10.3% (10.1%-10.5%) [14] |
|  | Delta, non-household | 3.0% (2.8%-3.2%) [14] |
|  | Omicron, household | 15.8% (14.3%-17.5%) [14] |
|  | Omicron, non-household | 8.7% (7.5%-10.0%) [14] |
| Vaccine effectiveness against infection with the Delta SARS-CoV-2 variant | AZ1 (received 1 dose of the Oxford AstraZeneca vaccine) | 30% |
|  | AZ2 (received 2 doses of the Oxford AstraZeneca vaccine) | 60% [15] |
|  | PF1 (received 1 dose of the Pfizer vaccine) | 30% |
|  | PF2 (received 2 doses of the Pfizer vaccine) | 80% [15] |
|  | Booster (received any combination of three vaccine doses) | 60% [15] |
|  | Natural immunity (with SARS-CoV-2, no vaccination) | 50% |
|  | Vaccinated and natural immunity (any combination of vaccinations and infection with SARS-CoV-2) | 50% |
|  | Unprotected (unvaccinated and no prior SARS-CoV-2 infection) | 0% |
| Vaccine associated reduction in transmission of the Delta SARS-CoV-2 variant | AZ1 | 45% [33] |
|  | AZ2 | 70% [15] |
|  | PF1 | 45% [33] |
|  | PF2 | 84% [15] |
|  | Booster | 90% [15] |
|  | Natural immunity | 65% |
|  | Vaccinated and natural immunity | 65% |
|  | Unprotected | 0% |
| Vaccine associated reduction in severe disease (hospital admission and death) due to the Delta SARS-CoV-2 variant | AZ1 | 80% |
|  | AZ2 | 94% [15] |
|  | PF1 | 85% |
|  | PF2 | 97% [15] |
|  | Booster | 96% [15] |
|  | Natural immunity | 94% |
|  | Vaccinated and natural immunity | 99.5% |
|  | Unprotected | 0% |
| Baseline mortality rate (per 1,000 SARS-CoV-2 infections in the absence of vaccination) | 5-year age groups {0-4, 5-9, 10-14, 15-17, 18-19, 20-24, 25-29, 30-34, 35-39, 40-44, 45-49, 50-54, 55-59, 60-64, 65-69, 70-74, 75-79, 80+} | {0.0, 0.0, 0.0, 0.2, 0.3, 0.4, 0.8, 1.2, 2, 2, 4, 10, 16, 34, 54, 86, 124, 192, 192, 192} [34] |
| Hospital admission rate (per 1,000 SARS-CoV-2 infections in the absence of vaccination) | 5-year age groups {0-4, 5-9, 10-14, 15-17, 18-19, 20-24, 25-29, 30-34, 35-39, 40-44, 45-49, 50-54, 55-59, 60-64, 65-69, 70-74, 75-79, 80+} | {1 1 1 1 2 5 10 16 23 29 39 58 72 102 117 146 177 180 200 200} [34] |
| Lateral flow sensitivity |  | 50% [27] |
| Contact tracing effectiveness, *CTF* |  | 25% [26] |
| Face mask effectiveness |  | 25% [29] |

## Results

### Risk-mitigation behaviour

The ALSPAC and TwinsUK/CSS Biobank surveys received 2,686 and 6,155 responses respectively. Most respondents (78% and 88% respectively) were aged between 30 and 69 years of age (Table 2). The vast majority of respondents to the ALSPAC survey (96%) were ALSPAC participants.

**Table 2:** Number of responses to the 2021 Christmas survey run by ALSPAC and TwinsUK/CSS Biobank.

| Age group | ALSPAC | Twins UK/CSS Biobank |
| --- | --- | --- |
| Under 30 | 406 | 221 |
| 30-39 | 231 | 1,348 |
| 40-49 | 22 | 1,338 |
| 50-59 | 592 | 1,348 |
| 60-69 | 821 | 1,368 |
| 70-79 | 56 | 476 |
| 80 + | 0 | 52 |
| Prefer not to say | 0 | 4 |

The general patterns of risk mitigation measures were similar in both surveys, although TwinsUK/CSS Biobank participants reported slightly higher levels of risk mitigations than ALSPAC participants (Fig 1A & 1C). In both surveys, a high proportion of respondents (over 95%) reported some risk mitigation behaviours. The most frequently reported was getting vaccinated or boosted. Overall, 94% of ALSPAC and 97% of TwinsUK/CSS Biobank participants reported that they had or planned to get fully vaccinated, with minimal differences between age groups. Using home testing kits was second to vaccination: 78% of ALSPAC participants and 88% of TwinsUK/CSS Biobank participants planned to use lateral flow testing kits before meeting friends and family. There was a decline in the use of home testing kits with age in the TwinsUK/CSS Biobank data.

**Figure 1:**
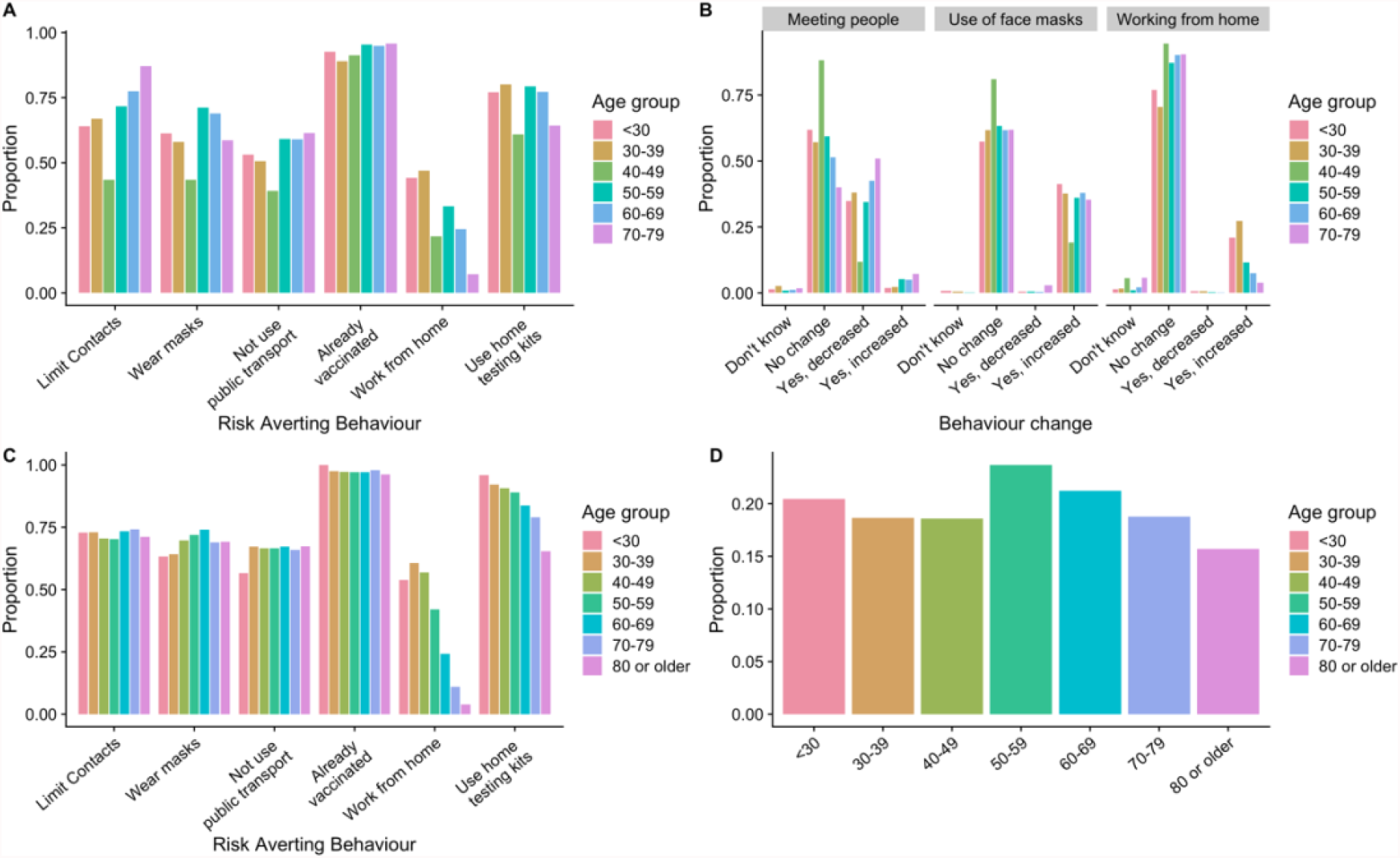
Survey responses from ALSPAC (Avon Longitudinal Survey of Parents and Children) and TwinsUK/CSS Biobank. (A): The proportion of ALSPAC respondents (N=2,686) by age group reporting risk mitigation measures during the period 20 December 2021 to 2 January 2022 inclusive. (B): The proportion of ALSPAC respondents who changed their behaviour (meeting people, use of face masks, working from home) due to the announcement of “plan B”. (C): The proportion of TwinsUK/CSS Biobank respondents (N=6,155) by age group reporting risk mitigation measures during the period 20 December 2021 to 2 January 2022 inclusive. (D): The proportion of TwinsUK/CSS Biobank respondents who changed their behaviour due to the announcement of “plan B”.

Face mask use was reported by 67% of ALSPAC and 70% of TwinsUK/CSS Biobank participants (Figures 1A and C). Of the respondents that reported planning to use a face mask, 64% said that “plan B” had no impact on face mask use, and 36% said that use would increase due to “plan B” (Fig 1B). In both surveys, 72% of respondents reported planning to limit contacts; nearly half of ALSPAC respondents who reported this said it had been affected by the announcement of “plan B”. Up to 25% of respondents in the TwinsUK/CSS Biobank survey reported altering their behaviour due to “plan B”, with the highest levels of change reported in 50-59 year olds (Fig. 1D).

We found that participants tended to report multiple risk mitigating measures. For example, being vaccinated, using face masks and limiting exposure was highly predictive of using home testing kits. The exception was working from home, which was not predictive of other risk mitigation measures.

### Omicron SARS-CoV-2 variant in England

Using the individual-based model, we estimate that the observed increase in secondary attack rates and measured reduction in vaccine effectiveness is consistent with an increase the effective reproduction number in England from unity to 2.9 (95%CI 2.5, 3.5) (Fig 2A). This estimate is essentially unaffected by assumptions about disease severity or vaccine effectiveness against severe disease (Figs 2D&2G).

**Figure 2:**
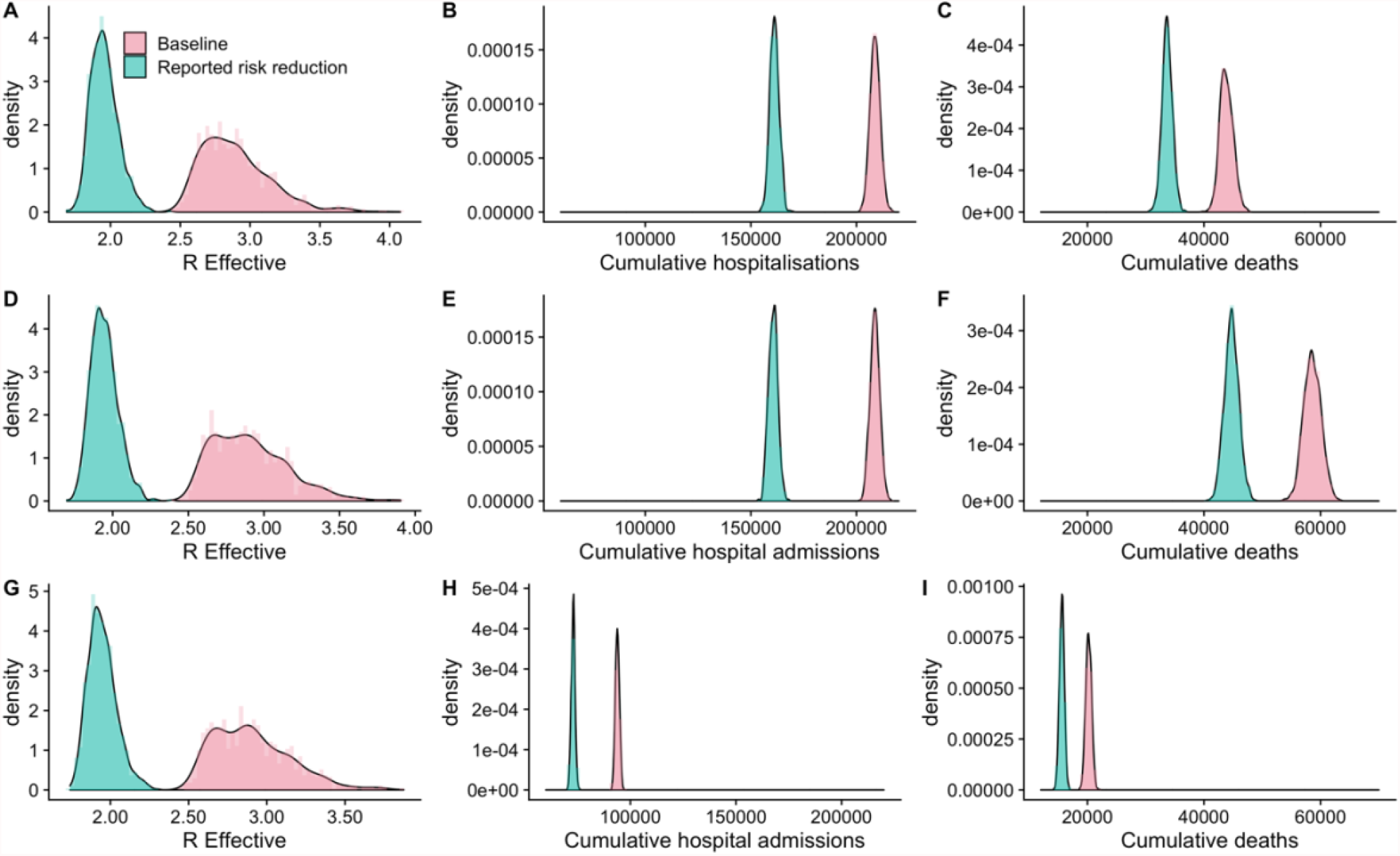
The estimated size of the effective reproduction number (panels A, D, G), cumulative hospital admissions (panels B, E, H) and cumulative deaths (panels C, F, I) with and without reported risk mitigation measures. Panels A, B, C: with a 40% reduction in severity associated with Omicron relative to Delta. Panels D, E, F: with a 20% reduction in severity associated with Omicron relative to Delta. Panels G, H, I: with a 20% reduction in severity associated with Omicron relative to Delta and assuming that vaccine effectiveness against severe disease is not reduced with Omicron infection. Vaccine distribution as of 26 November 2021.

In an unconstrained epidemic, this reproduction number translates to approximately 35 (95%CI 34, 36) million infections. The impact on hospital admissions and deaths is dependent on the relative severity of the Omicron variants compared to the Delta variant. Baseline with 40% reduction in severity relative to the Delta variant leads to an estimated 225,000 (95%CI 216,000 – 236,000) hospital admissions and 44,000 (95%CI 42,000 – 46,000) deaths (Fig 2B & 2C). With a 20% reduction in severity relative to the Delta variant, these numbers would be higher: 59,000 (95%CI 56,000 – 62,000) deaths (Fig 2D-2F). With a 20% reduction in severity relative to the Delta variant, and no reduction in vaccine effectiveness against severe disease, the estimated cumulative hospital admissions is 110,000 (95%CI 102,000 – 120,000) and the estimated cumulative number of deaths 20,000 (95%CI 19,000 – 21,000) (Fig 2G-2I).

### Impact of voluntary risk mitigation behaviours on the Omicron SARS-CoV-2 variant epidemic in England

With the high reported risk mitigation measures, seen across two cohort studies, we estimate that the Omicron effective reproduction number in England could be reduced to 1.9 (95%CI 1.8 – 2.2). This reduced reproduction number equates to a 25% reduction in the number of infections to approximately 26 (95%CI 25 - 27) million cumulative infections, and a similar reduction in cumulative deaths to 34,000 (95%CI 32,000 – 35,000) (Fig 2C), under the assumption of 40% reduction in both severity of Omicron relative to Delta and 40% reduction in vaccine effectiveness against severe disease relative to Delta.

Although relative severity is central to the absolute numbers of hospital admissions and deaths, the relative reduction due to risk mitigation behaviours is consistently between 20% and 25%.

Figure 3 illustrates how the effective reproduction number and cumulative number of deaths depend on the percentage of normal work and leisure contacts that take place. Without additional risk mitigation behaviours, over half of work and leisure contacts would need to be prevented to achieve a reproduction number of less than 1.

**Figure 3:**
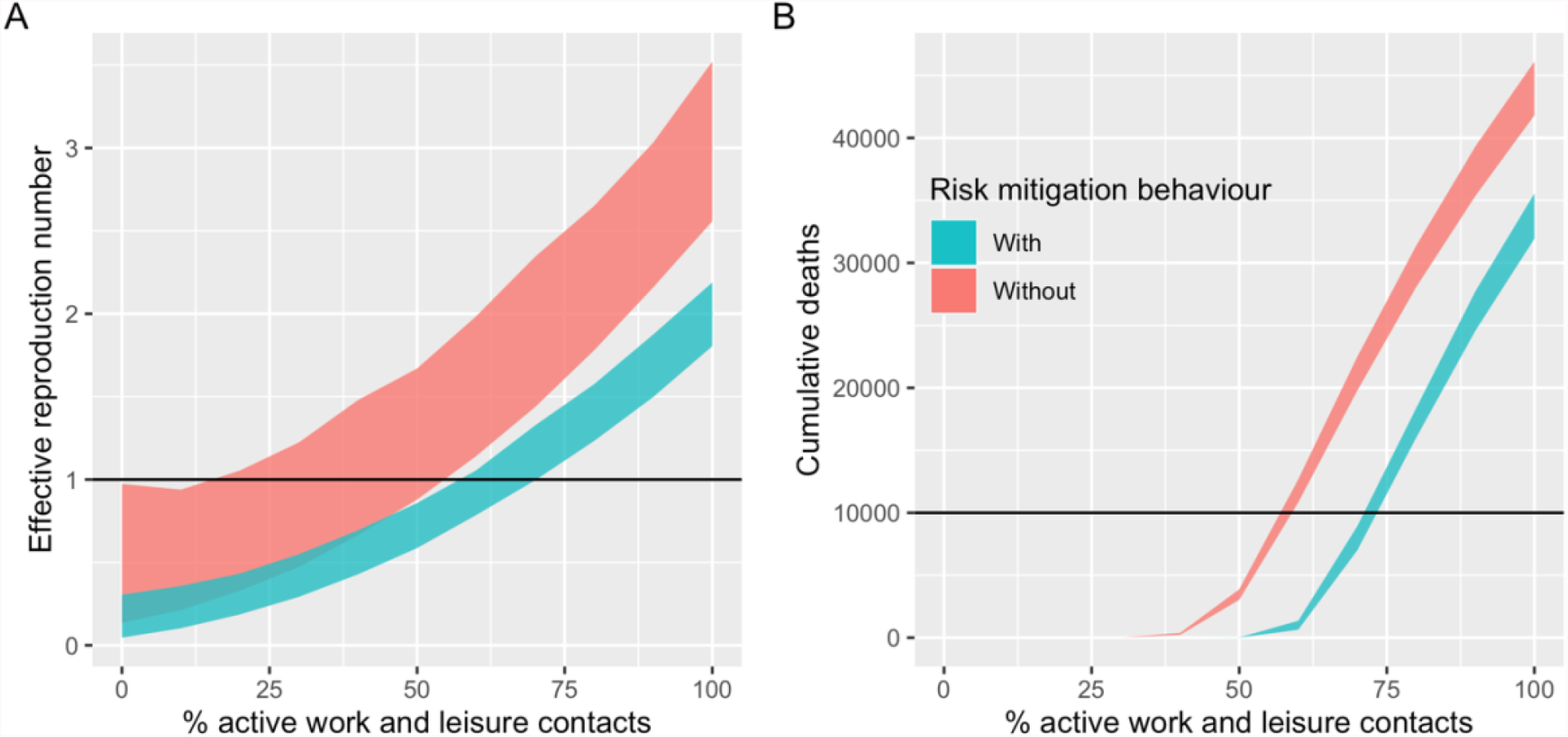
A: The effective reproduction number with and without risk mitigation behaviour. B: The cumulative number of deaths with and without risk mitigation behaviour.

## Discussion

Data here suggest that a high proportion of people intended to practice risk-mitigation behaviours to prevent the spread of COVID-19 during the Christmas 2021 period (from 20 December 2021 to 2 January 2022), with approximately 80% reporting using home testing kits. We estimate that these realistic risk-mitigating behaviours, including mask-wearing and regular home testing, could lead to a 40% reduction in the Omicron effective reproduction number and a 28% reduction in cumulative numbers of deaths. Nonetheless, these intended risk-mitigation behaviours are not sufficient to reduce the effective reproduction number below one, suggesting that epidemic control of Omicron cannot rely solely on self-imposed restrictions, and a further reduction in social contacts of 25-50% is required for control. Survey results also suggest that announcements of ‘Plan B’ restrictions had limited behavioural impact in our sample and would therefore have limited epidemiological impact.

Our results are based on reported intended behaviours, which might not reflect realised behaviours, for example due to additional information about the rapidly increasing number of COVID-19 cases, which may lead to increased awareness.. In order to rapidly gather behavioural information, we used established longitudinal cohorts which are designed to be broadly representative but nevertheless suffer from respondent bias with over-representation of females and higher socioeconomic status[18]. Our surveys did not include many children or young adults, and ALSPAC participants are mostly resident around Bristol. There are likely to be additional unmeasured biases in the surveys and understanding how risk-mitigation behaviour varies by age, ethnicity and socioeconomic status would be valuable for improved characterisation of the epidemiology in different communities.

There are other limitations in our modelling approach. As the model is not dynamic, we estimate the reproduction number and cumulative number of cases and deaths but cannot estimate the timescale over which cases and deaths occur or the peak burden of infection or deaths. The estimates of cumulative infections, hospital admissions and deaths are based on theoretical results that link the reproduction number to cumulative burden over the entirety of an epidemic. That means that this method intrinsically assumes an unmitigated epidemic, with no changes in behaviour, policy, adherence to guidelines or biological properties of COVID-19. In reality of course, epidemics are rarely unmitigated, especially if they are large.

This simplified modelling framework was used to inform decision-making in real-time. Our approach is less computationally intensive than dynamic models and by using theoretical results we provide a fast, transparent, and intuitive understanding of how behaviour translates to hospital admissions and deaths. In contrast to the majority of dynamic models used for forward epidemic projections [30–32], our approach is individual-based where individuals are explicitly modelled and scaled up to the population-level. This means that individual-level survey data can be readily incorporated, along with the associated uncertainties.

The main source of uncertainty in our modelled estimates is uncertainty in the severity of disease, both intrinsic severity (the probability of severe disease in a naïve, unvaccinated individual) and the probability of severe disease in individuals with prior immunity that suffer breakthrough infections. Under our baseline scenario, we estimated approximately 50,000 further deaths due to Omicron alone, which is a 30% increase in the total number of deaths in England between March 2020 and December 2021.

This study was used in real-time to quantify the necessity of additional restrictions over-and-above voluntary measures. This work demonstrates the power of utilising existing longitudinal cohorts for rapid elicitation of behavioural information in combination with population-levels transmission models. Fostering such links improves the accuracy and utility of disease models used for public health action and decision making especially for rapidly spreading infections.

## Contributions

EBP, KN, NJT and LD were involved in the conceptualisation of the study, including the formulation or evolution of overarching research goals and aims. EBP, LP, FC, and LD performed the analysis, including statistical analysis, creation and interpretation of mathematical models. ACT, EN, DAM informed the immunological parameters of the study. EBP and LD prepared the initial manuscipt and created the work and figures. KN, NJT, CS, VB and MPG collected, processed, cleaned, and maintained study data and obtained ethics approval. All authors had access to the underlying data.

## Data Availability

Aggregate data are available with this paper. Access to anonymous individual-level data can be applied for in line with the ALSPAC data access policy at http://www.bristol.ac.uk/alspac and/or the TwinsUK website https://twinsuk.ac.uk/resources-for-researchers/access-our-data/.

## Acknowledgements

We are grateful to everyone who took part in this study, the ALSPAC team (including interviewers, computer and laboratory technicians, clerical workers, research scientists, volunteers, managers, receptionists and nurses). Thank you to the TwinsUK and CSS Biobank teams, including the Volunteer Advisory Panels for their review and input to refine the survey.

ALSPAC and TwinsUK are both part of the National Core Studies, an initiative funded by UKRI, NIHR and the Health and Safety Executive. The COVID-19 Longitudinal Health and Wellbeing National Core Study was funded by the Medical Research Council (MC_PC_20030). Data collection efforts deployed as for this work form part of the “NCS” programme and were made possible because of the tireless dedication, commitment and enthusiasm of the many people who have taken part. We would like to thank the participants and the numerous team members involved in the studies including interviewers, technicians, researchers, administrators, managers, health professionals and volunteers.

---

**Supplementary Table 1:** The percentage of individuals by age group by vaccine and immune status, assuming 50% case ascertainment..

| Age group | N | AZ1 | AZ2 | PF1 | PF2 | MOD1 | MOD2 | Booster | Any vaccination | Unvaccinated | Test positive | Natural Immunity only (no vaccination) | Unprotected |
| --- | --- | --- | --- | --- | --- | --- | --- | --- | --- | --- | --- | --- | --- |
| 0-4 | 3,299,637 | 0.0 | 0.0 | 0.0 | 0.0 | 0.0 | 0.0 | 0.0 | 0.0 | 100.0 | 5.9 | 11.7 | 88.3 |
| 5-9 | 3,538,206 | 0.0 | 0.0 | 0.0 | 0.0 | 0.0 | 0.0 | 0.0 | 0.0 | 100.0 | 13.2 | 26.5 | 73.5 |
| 10-14 | 3,354,246 | 0.0 | 0.0 | 25.0 | 0.3 | 0.0 | 0.0 | 0.0 | 25.3 | 74.7 | 25.3 | 50.4 | 49.2 |
| 15-17 | 1,831,479 | 0.1 | 0.4 | 54.2 | 9.8 | 0.0 | 0.0 | 0.4 | 64.6 | 35.4 | 25.5 | 45.6 | 43.9 |
| 18-19 | 1,258,753 | 1.5 | 8.0 | 10.2 | 54.0 | 1.6 | 5.2 | 1.6 | 80.8 | 19.2 | 23.2 | 14.5 | 16.8 |
| 20-24 | 3,487,863 | 0.8 | 11.1 | 8.8 | 48.1 | 1.0 | 5.1 | 3.6 | 76.0 | 24.0 | 21.2 | 13.6 | 18.4 |
| 25-29 | 3,801,409 | 0.9 | 12.7 | 6.9 | 48.6 | 0.7 | 4.9 | 5.4 | 77.6 | 22.4 | 19.2 | 10.9 | 17.5 |
| 30-34 | 3,807,954 | 1.3 | 17.0 | 5.7 | 50.9 | 0.6 | 5.3 | 7.1 | 86.3 | 13.7 | 18.7 | 7.4 | 12.3 |
| 35-39 | 3,733,642 | 1.7 | 20.7 | 4.0 | 50.2 | 0.4 | 4.6 | 8.6 | 89.0 | 11.0 | 18.0 | 5.8 | 10.3 |
| 40-44 | 3,414,297 | 3.1 | 53.4 | 1.3 | 23.6 | 0.1 | 2.4 | 11.9 | 95.4 | 4.6 | 19.4 | 3.3 | 5.3 |
| 45-49 | 3,715,812 | 2.6 | 53.6 | 1.4 | 17.5 | 0.2 | 2.7 | 14.0 | 90.9 | 9.1 | 16.6 | 4.0 | 8.1 |
| 50-54 | 3,907,461 | 2.0 | 54.0 | 0.8 | 14.7 | 0 | 0.3 | 25.4 | 95.4 | 4.6 | 15.1 | 1.7 | 3.9 |
| 55-59 | 3,670,651 | 1.8 | 49.6 | 0.6 | 15.6 | 0.0 | 0.1 | 32.9 | 99.2 | 0.8 | 13.3 | 0.5 | 1.4 |
| 60-64 | 3,111,835 | 1.4 | 39.0 | 0.4 | 15.1 | 0.0 | 0.1 | 45.7 | 99.9 | 0.1 | 11.1 | 0.0 | 0.1 |
| 65-69 | 2,796,740 | 0.7 | 22.1 | 0.2 | 11.0 | 0.0 | 0.0 | 65.4 | 98.5 | 1.5 | 7.8 | 0.2 | 1.3 |
| 70-74 | 2,779,326 | 0.1 | 12.6 | 0.6 | 7.2 | 0.0 | 0.0 | 79.8 | 99.6 | 0.4 | 6.3 | 0.0 | 0.3 |
| 75-79 | 1,940,686 | 0 | 3.3 | 0.4 | 3.0 | 0.0 | 0.0 | 93.9 | 99.2 | 0.8 | 6.6 | 0.0 | 0.0 |
| 80-84 | 1,439,913 | 0 | 4.1 | 1.0 | 11.0 | 0.0 | 0.0 | 84.4 | 99.5 | 0.5 | 7.3 | 0.1 | 0.4 |
| 85-89 | 879,778 | 0.9 | 4.9 | 0 | 11.5 | 0.0 | 0.0 | 82.7 | 98.2 | 1.8 | 10.2 | 0.2 | 0.6 |
| 90+ | 517,273 | 1.4 | 9.7 | 0 | 13.1 | 0.0 | 0.0 | 75.0 | 97.8 | 2.2 | 15.5 | 0.7 | 1.5 |

## Appendix

**ALSPAC Christmas survey questions** “Children of the 90s at Christmas” **Making a difference to the pandemic – what are your plans for Christmas?**

For the questions, please think about the people you might be planning to spend time with <u>indoors</u> over the festive period. By this we mean **20**^**th**^ **December to 2**^**nd**^ **January inclusive**. This might include for example, visits to/from family members, get-togethers with friends at the pub or in a restaurant, an office party or other social event.

1. How many people <u>in total</u> in each of the following age groups do you think you will spend time with (for at least an hour) <u>indoors</u> over the festive period. options are: none, 1-4, 5-9, 10+, don’t know
  a. Pre-school children (aged under 5 years)
  b. School/college aged children (aged 5 to 17 years)
  c. Young Adults (aged 18-29 years)
  d. Adults (aged 30-59 years)
  e. Older adults (aged 60+ years)
2. How many other households, **excluding your own**, are you planning to spend time with (for at least an hour) <u>indoors</u> over the festive period?
  f None (I will stay with my household only)
  g 1
  h 2
  i 3
  j 4
  k 5 or more
  l. Don’t know
3. Will you be using any of the following precautionary measures during the festive period? Yes/No/Don’t know for each question
  a. Use home testing kits before meeting friends or relatives?
  b. Work from home in the days before meeting friends or relatives?
  c. Limit your contacts/exposure risk in the days before meeting friends or relatives?
  d. Get vaccinated / receive the booster vaccine
  e. Shop online, instead of visiting shops
  f. Not using public transport
  g. Increase indoor ventilation
  h. Wear a mask in indoor spaces
4. Have you and the current members of your household been vaccinated against COVID-19?
  a. Yes, all eligible household members have been vaccinated with at least one dose
  b. Some eligible household members have been vaccinated with at least one dose
  c. No household members have received a vaccination
  d. I don’t know
5. How old are you? < 30 30-39 40-49 50-59 60-69 70-79 80+ Prefer not to say
6. What is your gender? Male Female Non-Binary Prefer not to say
7. Please tell us the **first part** of your postcode e.g. if your postcode is BS1 9XX, please enter B in box 1, S in box 2 and 1 in box 3, leaving box 4 blank. If it is BS99 1XX, please enter B in box 1, S in box 2, 9 in box 3 and 9 in box 4. Box 1 Box 2 Box 3 Box 4
8. Where will you be working **most of the time** in the lead up to the festive period? Please only tick one box At Home Healthcare setting such as hospital, doctor’s surgery, care home Another setting where I will be in contact with other people (e.g. supermarket, office) Another setting where my contact with other people will be limited (e.g. lorry driving, Other
9. How will your patterns of activity change over the festive period compared to now? I will meet more people I will meet fewer people I will meet approximately the same number of people. I don’t know
10. Which generation of “Children of the 90s” study are you? Parent Original Child (born in 1990-1993) or partner of an original child I’m not a participant in the study

